# Large language models to help appeal denied radiotherapy services

**DOI:** 10.1101/2024.04.30.24306634

**Authors:** Kendall Kiser, Mike Waters, Jocelyn Reckford, Christopher Lundeberg, Christopher Abraham

## Abstract

**Background:** Large language model (LLM) artificial intelligences have potential to perform myriad healthcare tasks but should be validated in specific clinical use cases before deployment. One use case is to help physicians appeal insurer denials of prescribed medical services, a task that delays patient care and contributes to burnout. We evaluated LLM performance at this task for denials of radiotherapy services.

**Methods:** We evaluated generative pre-trained transformer 3.5 (GPT-3.5) (OpenAI, San Francisco, CA), GPT-4, GPT-4 with internet search functionality (GPT-4web), and GPT-3.5ft. The latter was developed by fine-tuning GPT-3.5 via an OpenAI application programming interface with 53 examples of appeal letters written by radiation oncologists. Twenty test prompts with simulated patient histories were programmatically presented to the LLMs, and output appeal letters were scored by three blinded radiation oncologists for language representation, clinical detail inclusion, clinical reasoning validity, literature citations, and overall readiness for insurer submission.

**Results:** Interobserver agreement between radiation oncologists’ scores was moderate or better for all domains (Cohen’s kappa coefficients: 0.41 – 0.91). GPT-3.5, GPT-4, and GPT-4web wrote letters that were on average linguistically clear, summarized provided clinical histories without confabulation, reasoned appropriately, and were scored useful to expedite the insurance appeal process. GPT-4 and GPT-4web letters demonstrated superior clinical reasoning and were readier for submission than GPT-3.5 letters (p < 0.001). Fine-tuning increased GPT-3.5ft confabulation and compromised performance compared to other LLMs across all domains (p < 0.001). All LLMs, including GPT-4web, were poor at supporting clinical assertions with existing, relevant, and appropriately cited primary literature.

**Conclusions:** When prompted appropriately, three commercially available LLMs drafted letters that physicians deemed would expedite appealing insurer denials of radiotherapy services. LLMs may decrease this task’s clerical workload on providers. However, LLM performance worsened when fine-tuned with a task-specific, small training dataset.

## Introduction

In the United States, medical insurers may deny reimbursement of some medical services they deem inappropriate for a patient.^1,2^ Patients’ physicians are permitted to appeal coverage denials through avenues the insurer stipulates, and which usually involve telephone conversations with insurer designees (“peer-to-peer” discussions) and formal appeal letters. Insurers assert that this practice adjudicates the best use of medical resources and protect patients from unnecessary medical interventions, but physicians across a spectrum of medical specialties argue that these practices delay patient care,^3,4^ discriminate,^5,6^ deter enrollment to and confound interpretation of clinical trials,^7,8^ and burden physicians^9-12^ with unnecessary clerical work and costs.^13^ Cancer patients – whose lives might depend on multidisciplinary systemic, surgical, and radiotherapeutic treatments – are too familiar with insurer delays and denials.^14,15^ In a cross-sectional survey, 22% of cancer patients reported that they did not receive care as originally recommended by their oncologists.^16^ At 97% of services, radiation oncology has been reported to be the medical specialty with the highest proportion of services for which insurers require authorization prior to treatment.^2^ One academic radiation oncology practice estimated its annual prior authorization-related cost burden to be nearly $500,000.^17^ Radiotherapy services that are initially denied and then appealed have been reported to be eventually authorized in 47%^18^ – 68%^19^ of instances.

Large language model (LLM) artificial intelligences have attained notoriety for their successes at interpreting and responding to human language,^20-22^ and LLM uses are emerging in healthcare. For example, in 2023 the New York University Langone Health System trained and fine-tuned an LLM (NYUTron) on unstructured text from 7.25 million clinical notes to perform clinical and operational tasks.^23^ In a prospective single arm trial, NYUTron predicted hospital readmissions from physician discharge summaries with an area of the curve (AUC) of 78.7%. Considering this and other successes, LLMs might be able to write suitable letters to appeal denied medical services.^24^ One case report detailed use of an LLM to obtain prior authorization for an orthopedic procedure,^25^ but to our knowledge no physician evaluation of LLM performance at this task has yet been reported, nor an attempt to fine-tune an LLM with training data suited to this task. In this study, we prompted three publicly accessible LLMs and one fine-tuned LLM to generate letters appealing denied radiotherapy services and we evaluated their outputs. We hypothesized that LLM performance at this task would be clinically useful and better still after fine-tuning.

## Methods

Formal letters written to appeal medical insurer decisions to deny coverage of radiotherapy services were collected from radiation oncologists at a single academic institution. A radiation oncologist reviewed the contents of all appeal letters and, for each letter, drafted a prompt matched to the letter’s content. Letters and paired prompts were used as training data to fine tune GPT-3.5 (OpenAI, San Francisco, CA), an LLM, in a Microsoft Azure workspace (Redmond, WA) that was compliant with Health Insurance Portability and Accountability Act requirements. After fine-tuning, a radiation oncologist prepared 20 test prompts, each of which requested an appeal letter output for a simulated clinical scenario (scenarios were written to be like those present in the training data). Ten test prompts were intentionally simplistic and provided minimal clinical background, while the other ten were clinically complex with complete simulated patient clinical histories (Supplementary Table 1). A subset of prompts requested that the output appeal letters reason with and cite primary literature sources.

Test prompts were programmatically presented to four LLMs: GPT-3.5, GPT-3.5 after fine-tuning (GPT3.5ft), GPT-4, and GPT-4 with internet search capability (GPT-4web). Three radiation oncologists, who were blinded to the LLM provenance, independently scored output letters according to a pre-specified rubric with the following domains: syntactic and semantic language representation, inclusion of prompt clinical details, validity of clinical reasoning, and overall readiness to submit to a medical insurer (Table 1). Additionally, in letters that referenced primary literature to support their claims, one radiation oncologist investigated whether the citations existed and scored them for accuracy and clinical relevance. Inter-observer scoring variability was evaluated with weighted Cohen’s Kappa coefficients. Kruskal-Wallis nonparametric tests were computed between LLM model scores for each rubric domain, and subsequent Mann-Whitney U tests were computed between pairs of LLM model score distributions for rubric domains with a statistically significant Kruskal-Wallis result. Where applicable for iterative statistical tests, a Bonferroni correction deflated the p value significance level. Statistics were computed using SciKit-Learn, Statsmodels, and SciPy Python statistical packages.

**Table 1:**
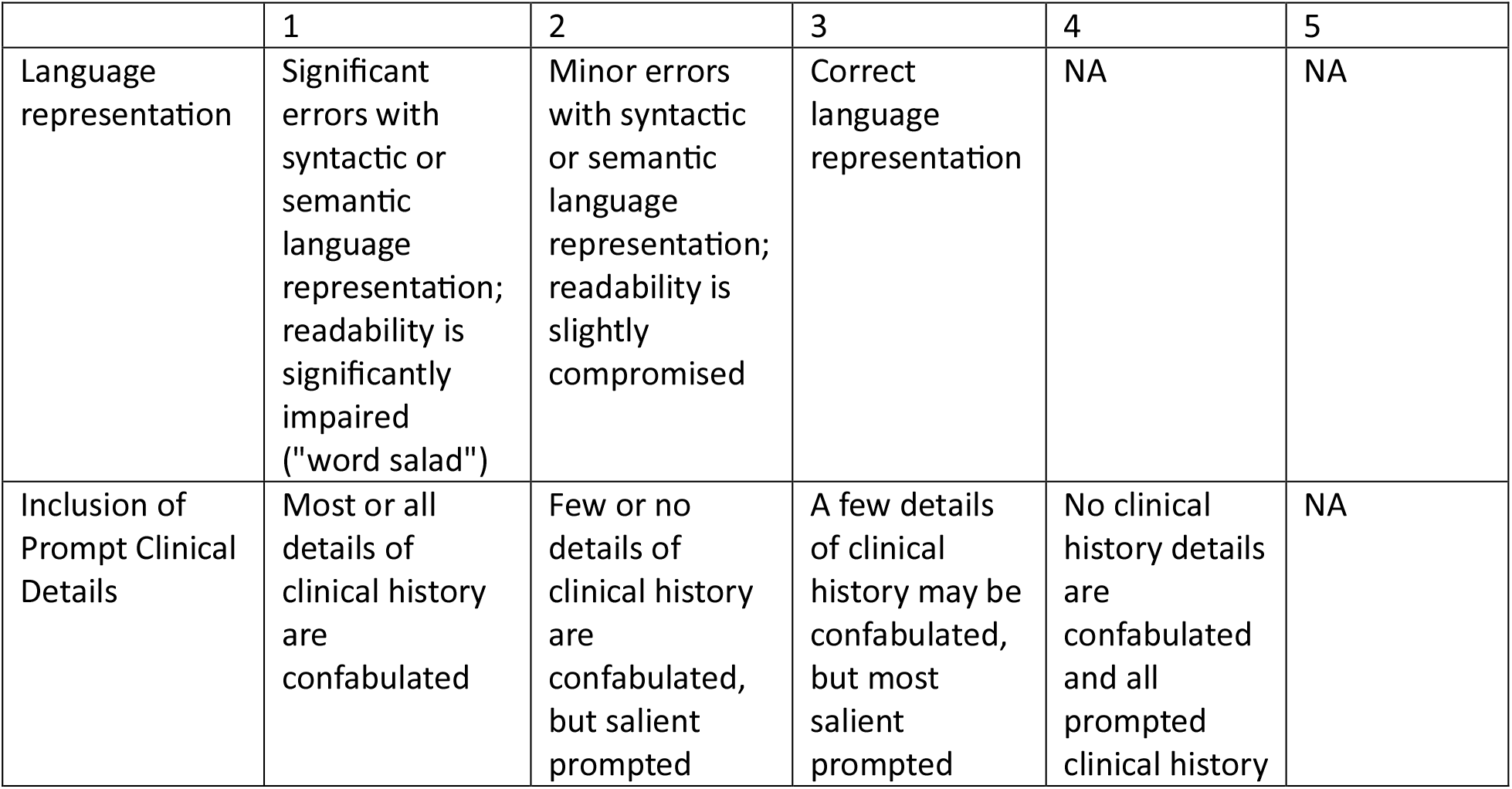
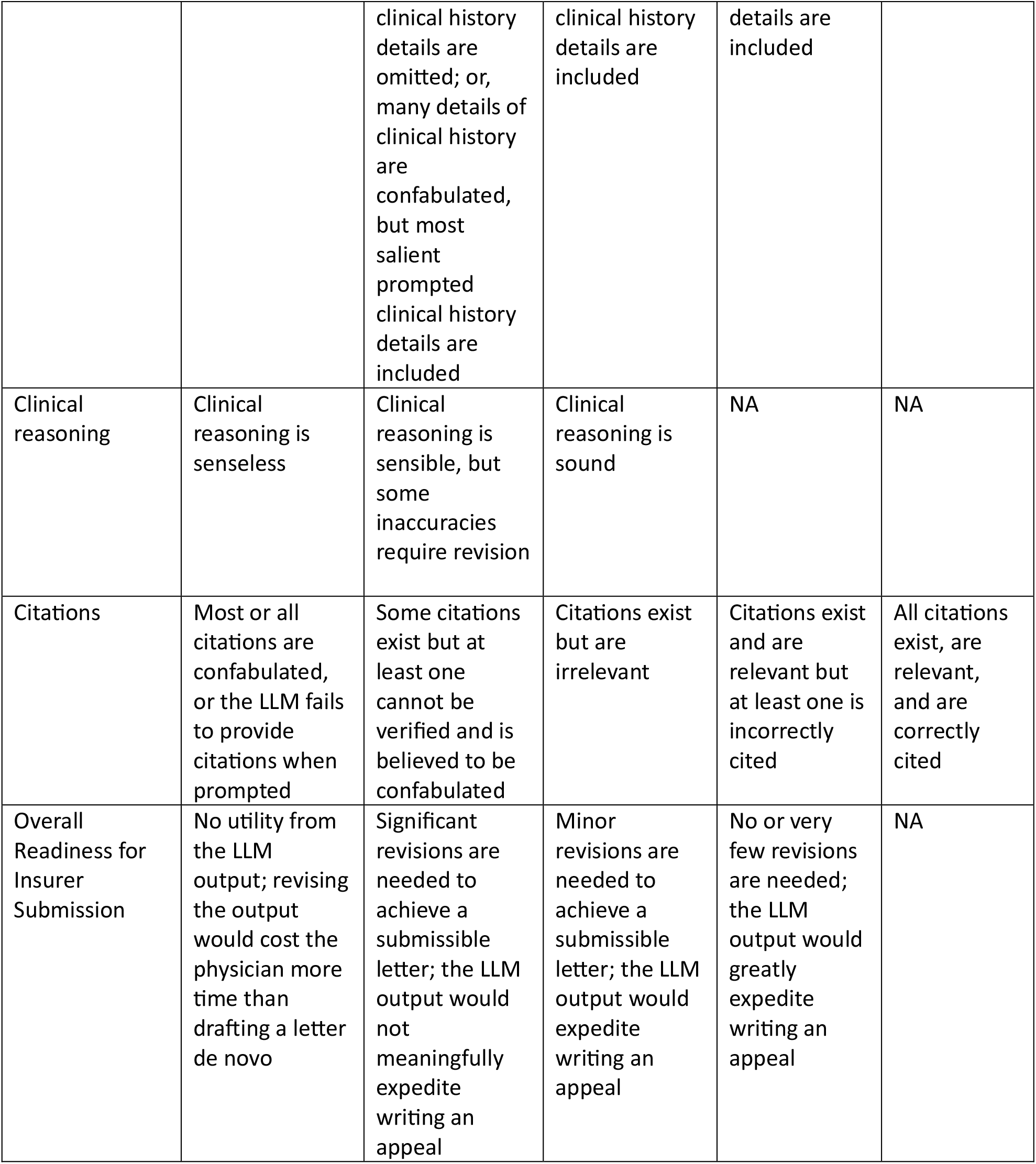
Scoring rubric for LLM output letters.

|  | 1 | 2 | 3 | 4 | 5 |
| --- | --- | --- | --- | --- | --- |
| Language representation | Significant errors with syntactic or semantic language representation; readability is significantly impaired ("word salad") | Minor errors with syntactic or semantic language representation; readability is slightly compromised | Correct language representation | NA | NA |
| Inclusion of Prompt Clinical Details | Most or all details of clinical history are confabulated | Few or no details of clinical history are confabulated, but salient prompted | A few details of clinical history may be confabulated, but most salient prompted | No clinical history details are confabulated and all prompted clinical history | NA |
|  |  | clinical history details are omitted; or, many details of clinical history are confabulated, but most salient prompted clinical history details are included | clinical history details are included | details are included |  |
| Clinical reasoning | Clinical reasoning is senseless | Clinical reasoning is sensible, but some inaccuracies require revision | Clinical reasoning is sound | NA | NA |
| Citations | Most or all citations are confabulated, or the LLM fails to provide citations when prompted | Some citations exist but at least one cannot be verified and is believed to be confabulated | Citations exist but are irrelevant | Citations exist and are relevant but at least one is incorrectly cited | All citations exist, are relevant, and are correctly cited |
| Overall Readiness for Insurer Submission | No utility from the LLM output; revising the output would cost the physician more time than drafting a letter de novo | Significant revisions are needed to achieve a submissible letter; the LLM output would not meaningfully expedite writing an appeal | Minor revisions are needed to achieve a submissible letter; the LLM output would expedite writing an appeal | No or very few revisions are needed; the LLM output would greatly expedite writing an appeal | NA |

## Results

Fifty-three letters that appealed radiotherapy coverage denials by medical insurers were collected for training data. The denied services included proton radiotherapy (n = 45), stereotactic ablative radiotherapy (n = 3), 3D conformal radiotherapy (n = 2), image-guided radiotherapy (n = 2), and intensity-modulated radiotherapy (n = 1). The clinical necessities for proton radiotherapy included re-irradiation of previously treated anatomic sites (n = 17; this was also the clinical necessity for one denial of intensity modulated radiation therapy, n = 1), inability to meet safe radiation dose tolerances for vulnerable organs with photon radiotherapy due to comorbid illnesses or atypical anatomic proximity to the radiation target (n = 13), young patient age (child or young adult; n = 5), history of connective tissue disease (n = 2), history of a germline cancer predisposition syndrome (n = 4), or enrollment on a national phase III clinical trial that randomized between proton and photon radiotherapy (n = 4). The clinical necessity for stereotactic ablative radiotherapy was treatment of oligometastatic or oligoprogressive cancer foci (n = 3). The clinical necessity for image-guided radiotherapy was image verification of correct radiotherapy target alignment adjacent to radiosensitive healthy tissues (n = 2). Two letters were submitted to clarify diagnostic codes and patient clinical history.

Eighty output insurance appeal letters – 20 per LLM – were independently scored by three blinded radiation oncologists. Score interobserver agreement was moderate-to-excellent for letter language representation (κ = 0.54 - 0.91), strong for clinical detail inclusion (κ = 0.73 - 0.78), moderate for clinical reasoning (κ = 0.41 - 0.62), and moderate-to-strong for overall readiness for submission to an insurer (κ = 0.67 - 0.77). LLM score distributions are visualized in Figure 1. The median language representation score was one for GPT-3.5ft and three for GPT-3.5, GPT-4, and GPT-4web. The median clinical detail inclusion score was one for GPT-3.5ft and four for GPT-3.5, GPT-4, and GPT-4web. The median clinical reasoning score was one for GPT-3.5ft, two for GPT-3.5, and three for GPT-4, and GPT-4web. The median overall submission readiness score was one for GPT-3.5ft and three for GPT-3.5, GPT-4, and GPT-4web. The median literature citation score was one for GPT-3.5ft, three for GPT-3.5 and GPT-4web, and three and one-half for GPT4.

**Figure 1:**
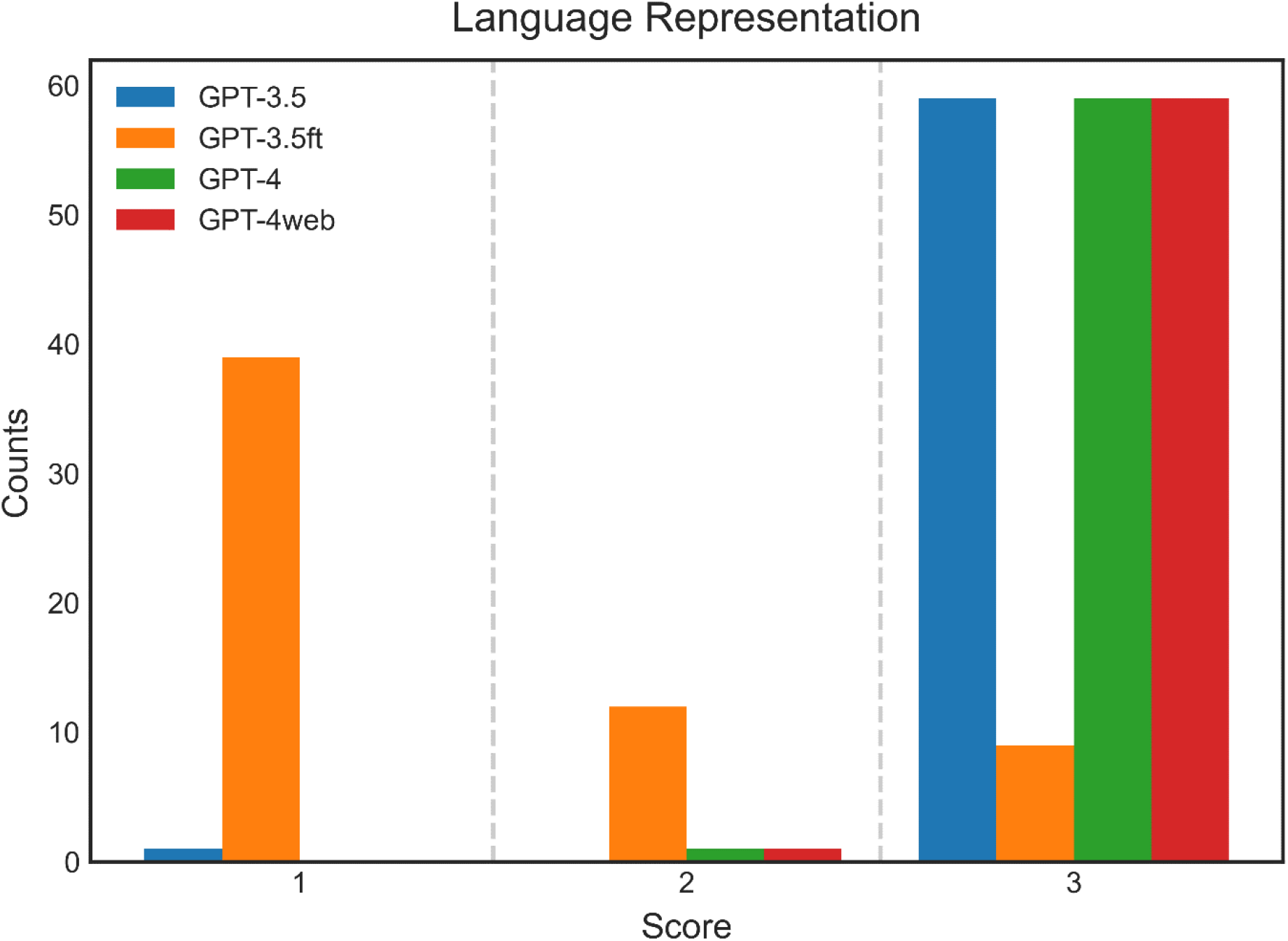
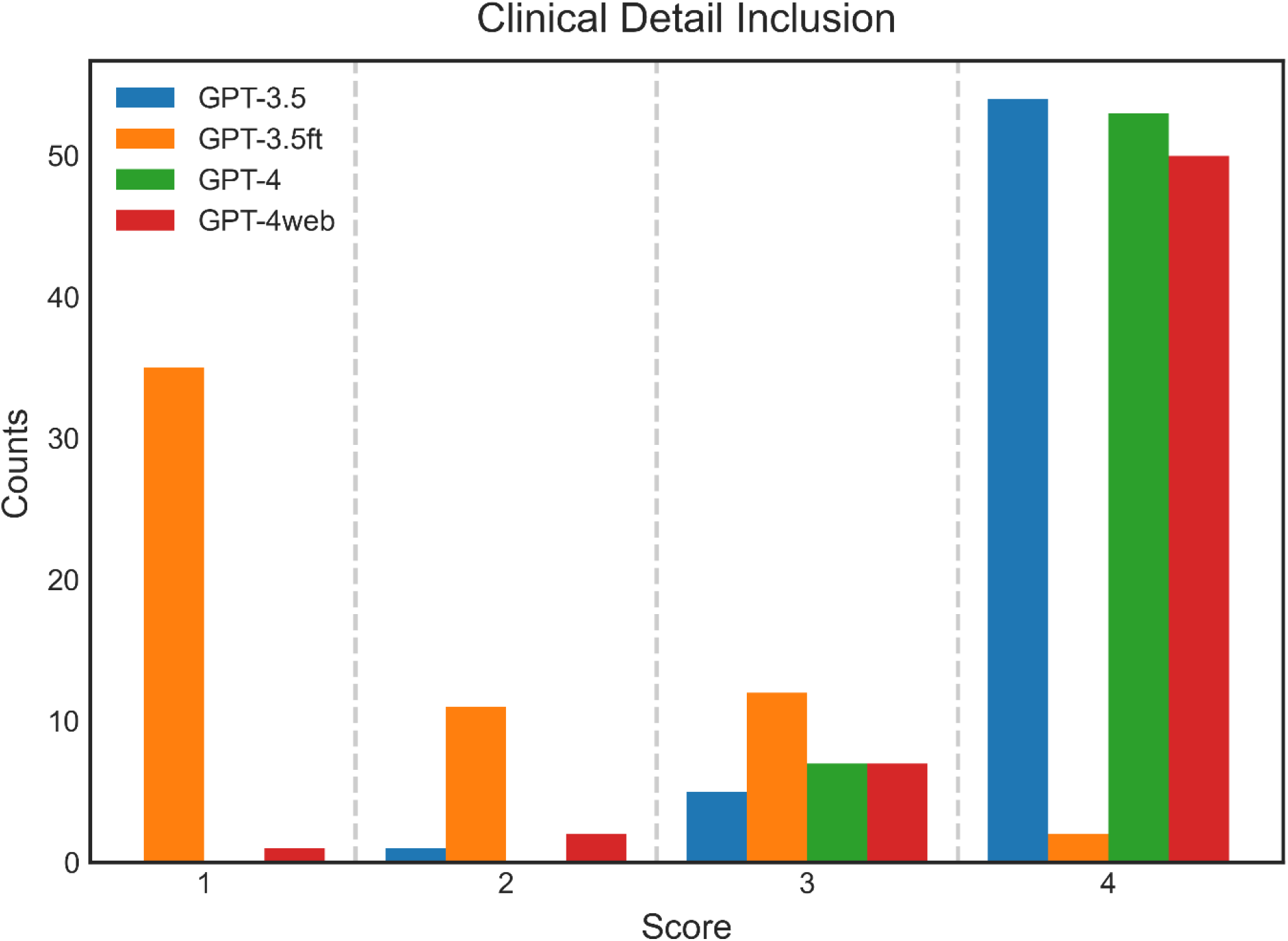
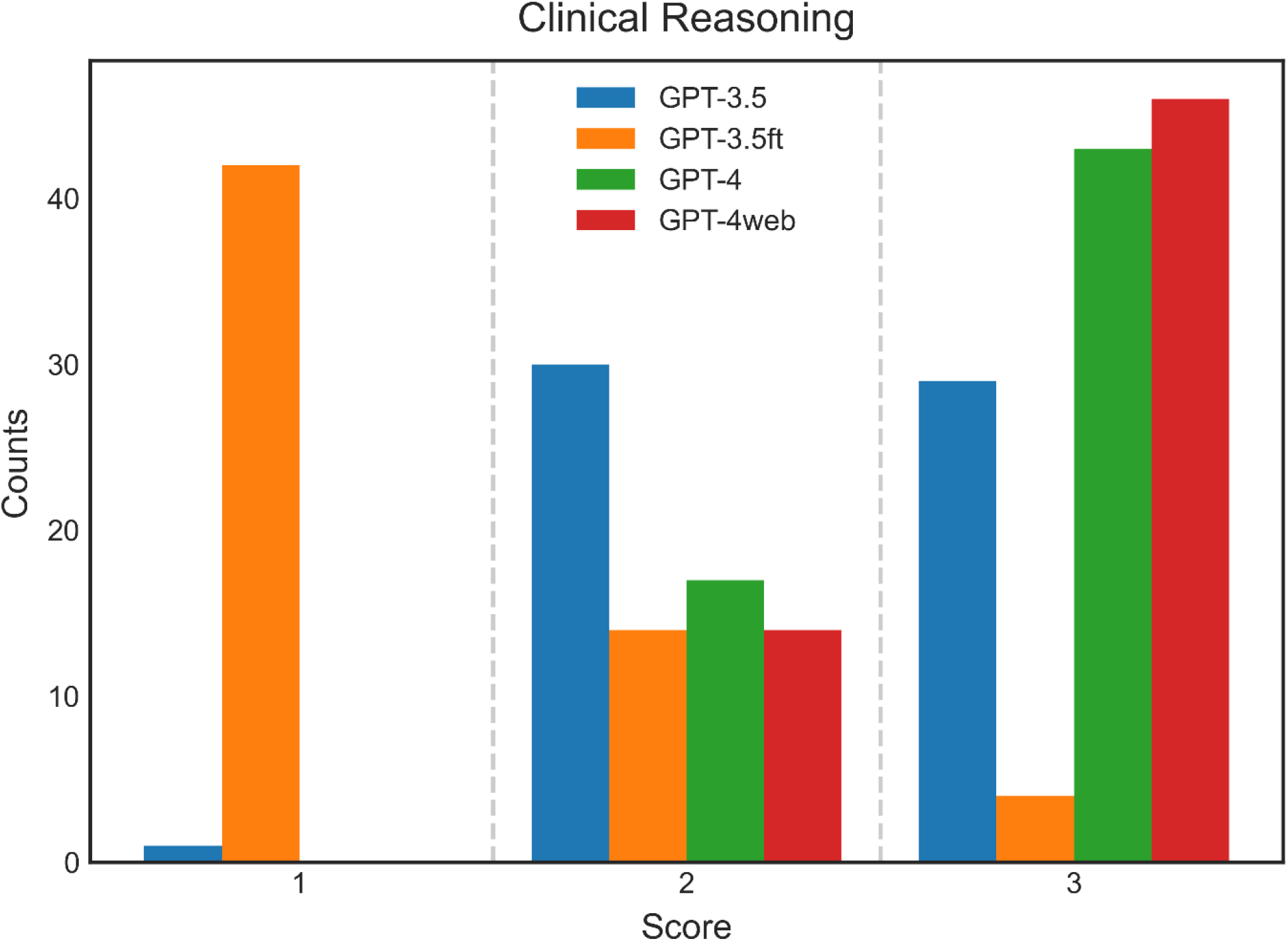
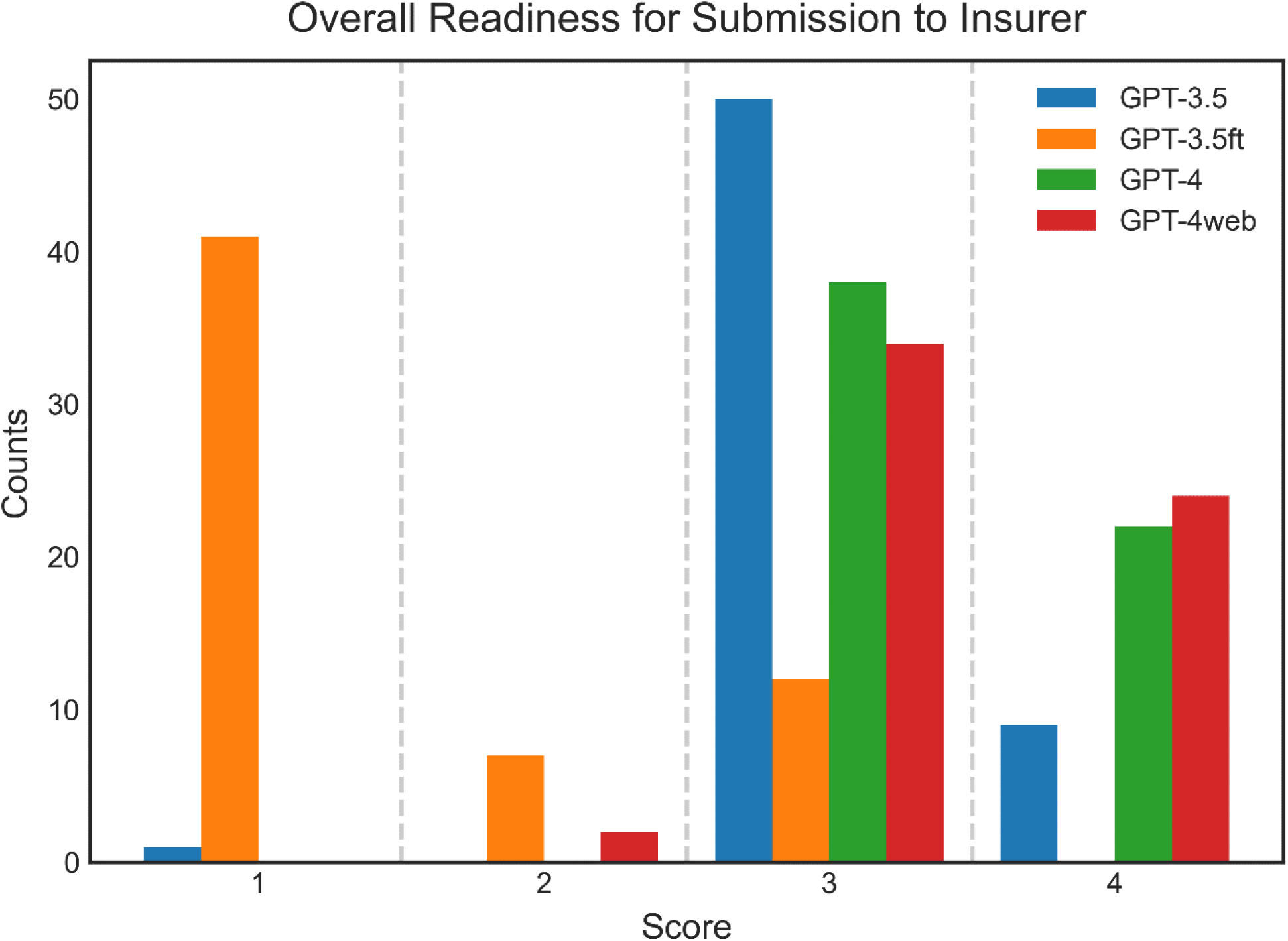
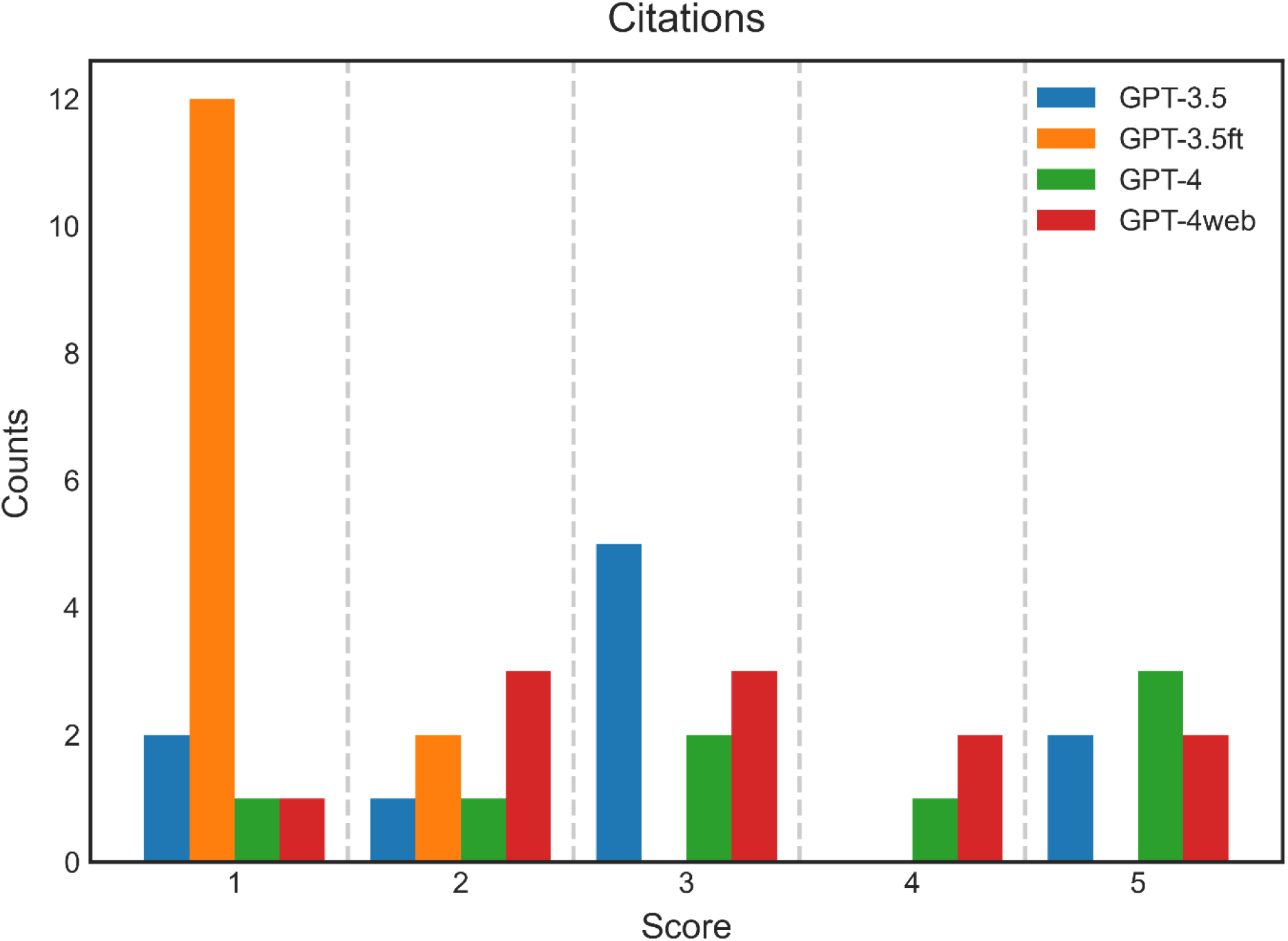
Frequency of LLM output letter scores for language representation (A), inclusion of prompt clinical detail (B), clinical reasoning (C), and overall readiness for insurer submission (D), as assessed by three radiation oncologists. Frequency of LLM output letter scores for primary literature citations (E), as assessed by one radiation oncologist.

Scores were not significantly different between GPT-3.5, GPT-4, and GPT-4web regarding language representation (p = 1.0) or prompt clinical detail inclusion (p = 0.49), but there were significant differences in elaboration of clinical reasoning (p = 0.002) and overall submission readiness (p = 0.008). Both GPT-4 and GPT-4web demonstrated enhanced clinical reasoning compared to GPT-3.5 (p = 0.008 and 0.001, respectively), but not compared to one another (p = 0.54). Likewise, letters produced by both GPT-4 and GPT-4web were deemed closer to ready for submission than letters produced by GPT-3.5 (p = 0.005 and p = 0.006, respectively), but readiness of letters produced by GPT-4 did not differ significantly from that of letters produced by GPT-4web (p = 0.90). Prompt complexity did not result in significantly different scores for most LLMs for most domains. Exceptions included GPT-4 readiness for submission (complex vs. simple mean scores: 3.50 vs. 3.23, p = 0.03) and GPT-4web clinical reasoning (complex vs. simple mean scores: 2.90 vs. 2.63, p = 0.02).

Fine-tuning significantly compromised GPT-3.5ft performance across all domains compared to the other LLMs (p < 0.001 for each domain) except for citing relevant primary literature (p = 0.80), which was challenging for all LLMs. GPT-3.5ft outputted letters that were on average twice as long as other LLMs (median word count 1183 vs. 600 (GPT-3), 603 (GPT-4), and 551 (GPT-4web); p values < 0.001) and often adopted language and letter structure used in the training data. Two qualitative patterns were observed in GPT-3.5ft outputs: 1) linguistic coherence deteriorated as the letter continued (Table 2), and 2) more frequent attempts to support assertions with literature citations were present – even without prompting – than in letters generated by other LLMs (Figure 1E), notwithstanding the citations’ poor accuracy and relevance. However, this observed difference did not achieve statistical significance (p = 0.80).

**Table 2:**
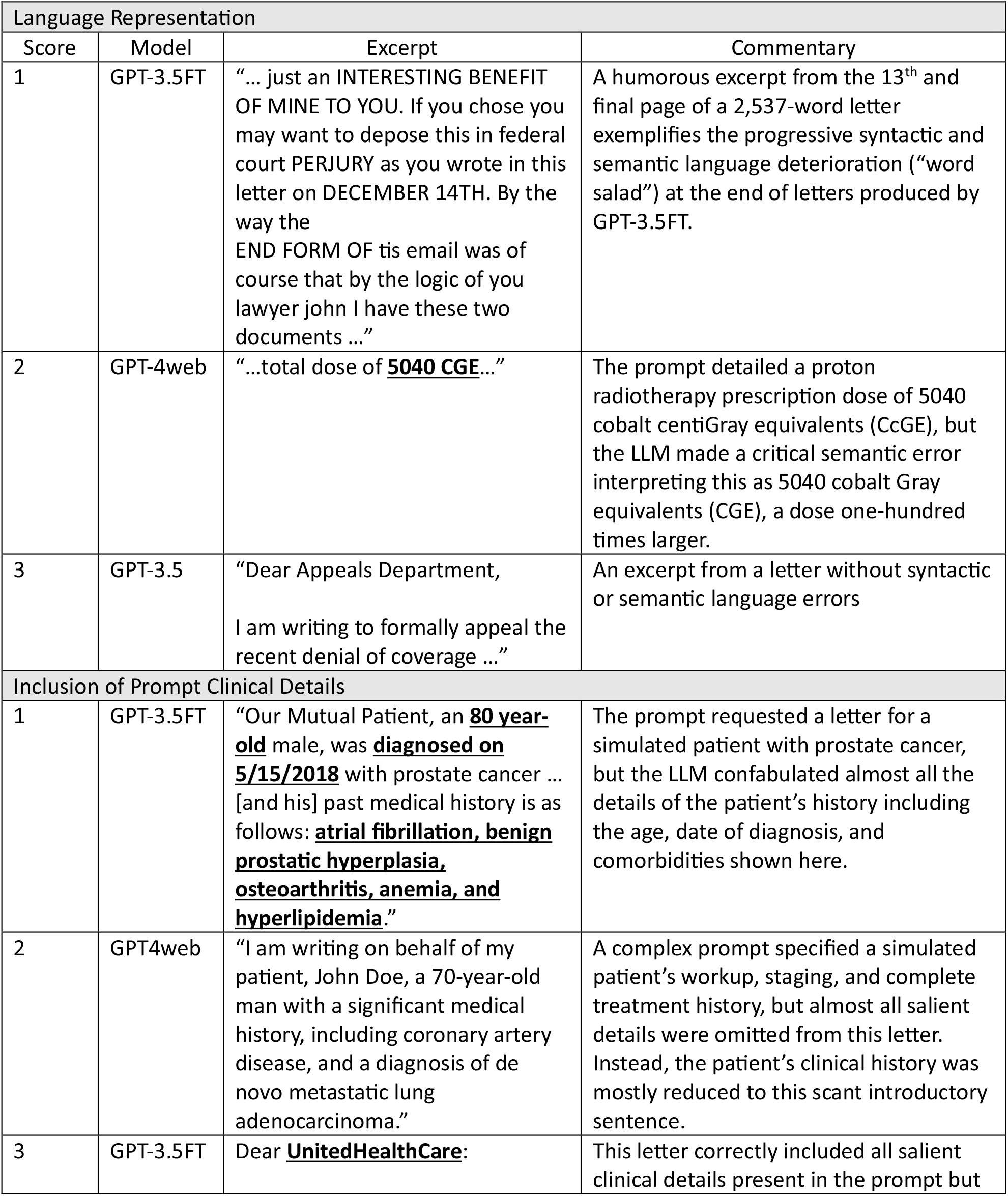
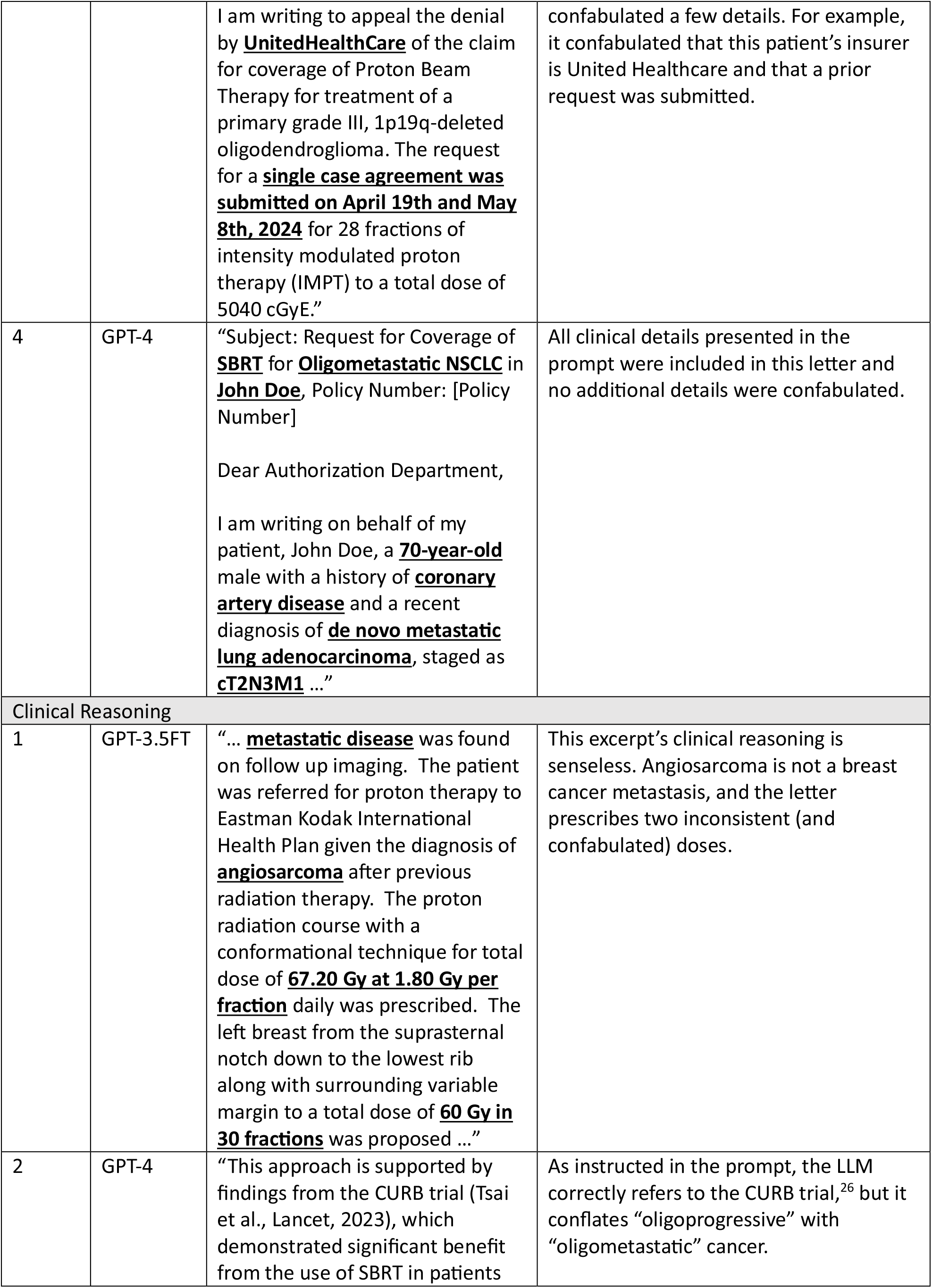
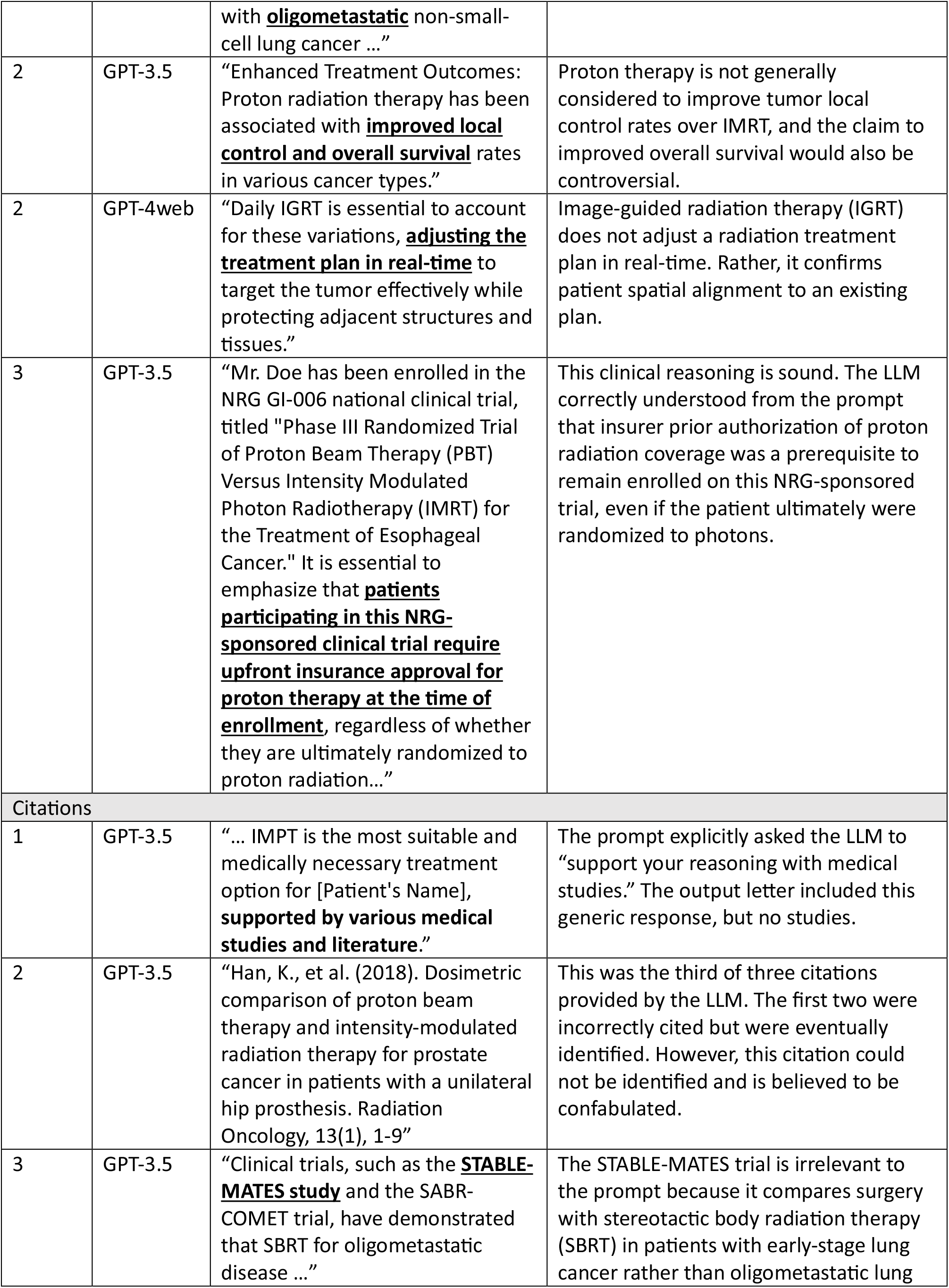
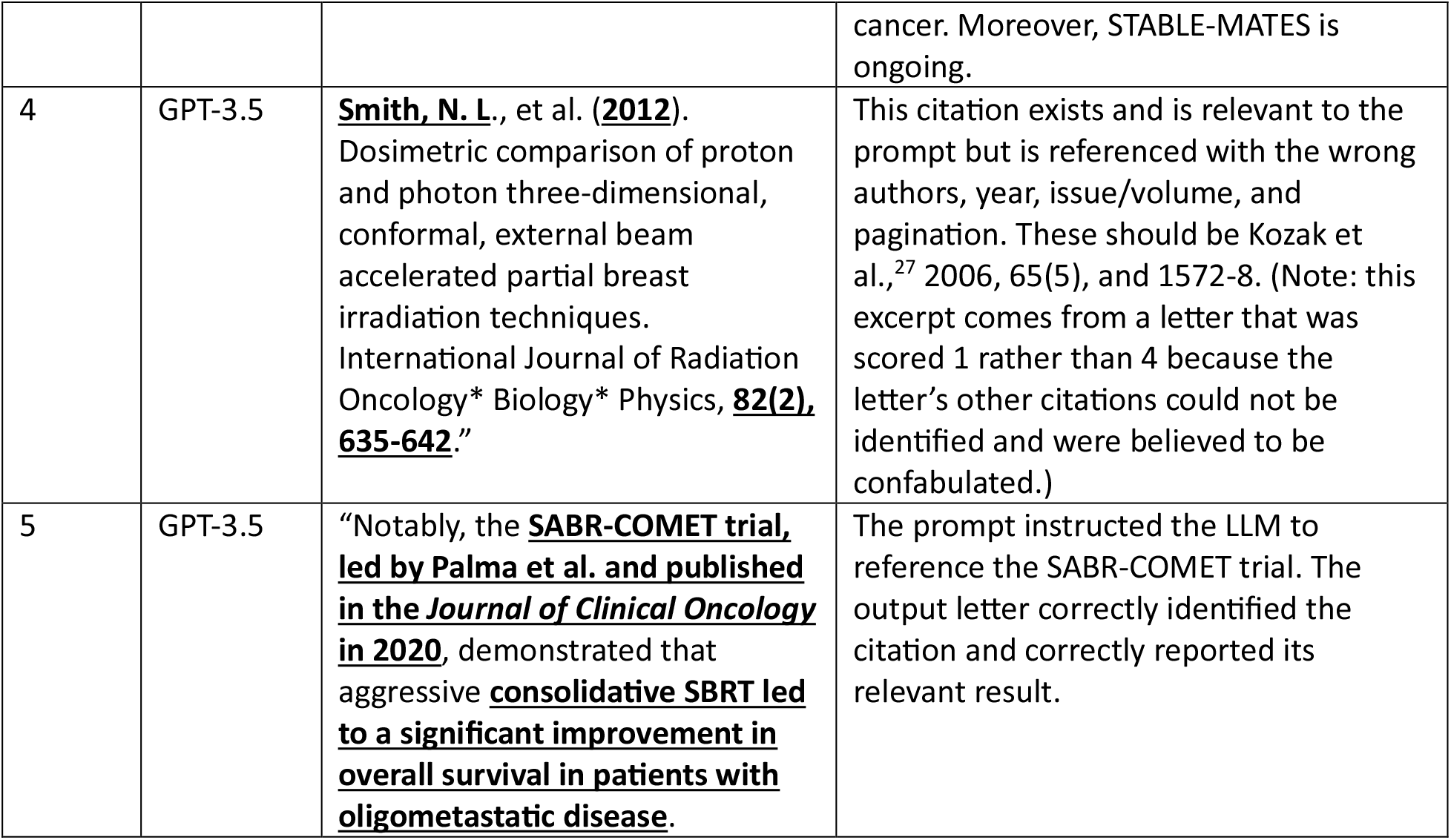
Excerpts from representative LLM output letters with associated scores across four domains.

## Discussion

GPT-3.5, GPT-4, and GPT-4web respectively produced letter drafts that 98%, 100%, and 97% of physician scores anticipated would expedite the clerical work required to appeal various denied radiotherapy services. These models responded with appropriate language, included salient prompt clinical details, and inferred with sound clinical reasoning in almost all prompted cases. Their responses were robust to test prompts that were developed with increased clinical complexity, and exploratory analysis suggested that GPT-4web and GPT-4web outputted letters that were significantly better reasoned and readier for submission with more complex prompts. Furthermore, GPT-4 and GPT-4web letters were significantly better reasoned and readier for submission than GPT-3.5 letters. By comparison, Katz et al. also reported that GPT-4’s clinical reasoning exceeded that of GPT-3.5 in the context of performance on Israeli residency board examinations.^28^ Importantly, all LLMs struggled to draft letters that incorporated support from relevant and appropriately cited primary literature sources. Empowering GPT4 with internet search functionality (as GPT-4web) did not appear to improve performance at this task. Overall, this study joins a nascent corpus of studies^23,28-33^ reporting physician validation of LLM performance at specific clinical tasks.

Contrary to our hypothesis, fine-tuning GPT-3.5 with 53 well-curated example letters worsened its performance, notwithstanding that the fine-tuned model began to adopt some patterns of language use and letter structure present in the training data. OpenAI instructs that improvements in GPT-3.5 performance can be seen after fine-tuning with as few as 50 training examples, but acknowledges that “the right number varies greatly based on the exact use case.”^34^ The developers of NYUTron similarly recommended “locally fine-tun[ing] an externally pre-trained language model when computation ability is limited.”^23^ However, the complexity of our clinical task clearly was greater than could be learned with a small number of training examples. Fine-tuning for our clinical task failed despite carefully curated training data, which were proofread first when submitted to a medical insurer and second when we formatted them for fine-tuning. Payne et al. fine-tuned GPT-4 with questions and answers from the American College of Radiology 2021 Diagnostic Radiology In-Training Examination (DXIT) but saw no improvement in GPT-4’s responses to the 2022 DXIT questions after fine-tuning.^31^ Our and Payne et al.’s results, while disappointing, are valuable because they suggest that fine-tuning may not be a solution for improving performance at all clinical tasks, particularly where training samples are limited. We do not know what number of training examples is necessary for an LLM to master nuances of writing insurance appeal letters, but we suspect that it is impractically high considering the impressive performance non-fine-tuned models already demonstrate.

This study’s strengths included the use of carefully reviewed training data, physician-curated prompts, and independent, blinded review of outputs by three radiation oncologists. Its limitations included a small number of training samples for the fine-tuning process and use of training data exclusive to radiotherapy appeals.

## Conclusions

Commercially available LLMs can incorporate complex clinical details and clinical reasoning into formal letters that are likely to expedite the clerical work physicians must complete to formally appeal denials of radiotherapy services. However, fine-tuning with task-specific training data made an LLM no better at this task, suggesting that fine-tuning may not always be a solution to improve LLM performance at specific clinical tasks.

## Data Availability

All data produced in the present study are available upon reasonable request to the authors

