## Supplemental Table 1 for "Large language models to help appeal denied radiotherapy services"

| Complexity | Appeal | Circumstance | Prompt |
|---|---|---|---|
| Simple | Proton | Left breast, young | Draft an official letter to appeal a medical insurer's denial of proton radiotherapy for the adjuvant treatment of a 30 year-old woman with a left breast pT2N1aM0 invasive ductal carcinoma s/p lumpectomy and sentinel lymph node dissection. She requires adjuvant proton RT because she is young. |
| Simple | Proton | Left breast, re-irradiation | I need a draft of a letter written to an insurance company to appeal its decision to deny proton radiation therapy to my patient, who is an elderly woman with a recurrent left breast invasive ductal carcinoma. She was diagnosed in 2023 with a left breast IDC s/p lumpectomy and adjuvant radiation therapy, and she now has a recurrent left breast invasive ductal carcinoma status post repeat lumpectomy. Explain that adjuvant radiation therapy with protons is indicated to protect her OARs in the context of re-irradiation. |

| | | | |
|---|---|---|---|
| Simple | Proton | Esophagus on trial (NRG GI-006) | Please provide me with a letter written to an insurance company that asks the insurer to approve reimbursement of a neoadjuvant proton radiation therapy course for a man with esophageal cancer who is on a national clinical trial, NRG GI-006. |
| Simple | Proton | Angiosarcoma | Write me a draft of a letter written to an medical insurance provider demanding it rescind denial of coverage for proton radiation therapy for my patient who has a left breast radiation-induced angiosarcoma. My patient requires proton (NOT photon) re-irradiation as part of the safe and efficacious treatment of her angiosarcoma. Please explain why in your letter. |

| | | | |
|---|---|---|---|
| Simple | Proton | Anus, young, Crohn's | I am a radiation oncologist, and I request that you draft an appeal letter written to a medical insurance company asking for coverage approval of proton radiation therapy for the definitive chemoradiation treatment of an anal cancer in a young man with locally advanced anal cancer, HIV, and Crohn's disease. The letter must convincingly argue that proton radiation is necessary, and that photon radiation, including IMRT, will not spare this young man's rectum and other organs as fully as proton radiation. |
| Simple | Proton | Brain, young | Please write a letter to an insurance provider appealing its decision to deny intensity modulated proton therapy (IMPT) as adjuvant treatment of a resected oligodendroglioma in a young male patient. Show how proton radiation therapy decreases long-term side effects of radiation compared to photon radiation, and support your reasoning with medical studies. |

| | | | |
|---|---|---|---|
| Simple | Proton | Generic | Please generate a generic letter template intended to appeal an insurance provider's rejection of medical coverage for proton radiotherapy. Reason why proton radiation therapy is in many cases dosimetrically superior to intensity modulated photon therapy. |
| Simple | SBRT | Oligometastatic lung | I need a letter written to an insurance provider that requests reimbursement/coverage for stereotactic body radiation therapy (SBRT) to a single right humeral metastasis in a patient with oligometastatic non-small-cell lung cancer. Please reason why stereotactic body radiation therapy can improve progression-free and overall survival in patients with oligometastatic disease, and reference the medical trials that support this claim. |

| | | | |
|---|---|---|---|
| Simple | SBRT | Oligoprogressive lung | I need a letter written to an insurance provider that requests reimbursement/coverage for stereotactic body radiation therapy (SBRT) to a single oligoprogressive right humeral metastasis in a patient with initially widely metastatic non-small-cell lung cancer whose metastatic disease is controlled except for this single, growing right humeral lesion. Please reason why stereotactic body radiation therapy can improve progression-free and overall survival in patients with oligoprogressive disease, and reference the medical trials that support this claim. |
| Simple | IGRT | 3D palliative re-irradiation | Explain to a medical insurer, in the form of a formal appeal letter, why daily image-guided radiation therapy must be covered and reimbursed to safely re-treat a sacral prostate cancer metastasis with palliative 3D radiation therapy one year after it was treated with prior palliative radiation therapy. |

| | | | |
|---|---|---|---|
| Complex | Proton | Left breast, young | Draft an official letter to appeal a medical insurer's denial of proton radiotherapy for the adjuvant treatment of Jane Doe, a 30-year-old woman with a left breast pT2N1aM0 invasive ductal carcinoma. I am this patient's treating radiation oncologist. Jane first presented to medical attention on January 1, 2024 after palpating a lump in her left breast. She underwent a diagnostic bilateral mammogram and left breast ultrasound. These visualized a 3.5 cm left breast mass in the upper outer quadrant at 2:00 a.m., 6 cm from the nipple. She underwent an ultrasound-guided biopsy on January 2, 2024. This demonstrated triple-negative, grade 3 IDC. On January 3, 2024 she completed 6 cycles of neoadjuvant chemoimmunotherapy. On January 4, 2024 she underwent a left modified radical mastectomy and right prophylactic skin sparing mastectomy with a left axillary nodal dissection. Pathology identified grade 3 invasive ductal carcinoma, pT2N1a, with 3.5 cm of residual primary disease and disease in 1 axillary lymph node, -SM, -LVSI, -ENE. She now requires adjuvant radiation therapy to her left chest wall and regional lymphatics to minimize the risk of locoregional recurrence. I have prescribed 42.56 cobalt Gy equivalents in 16 fractions. Please craft a letter justifying the use of proton radiation therapy, and why proton radiation therapy provides superior dosimetry to organs-at-risk compared to photon radiation therapy. Cite primary literature references to support your arguments. |
| Complex | Proton | Left breast, re-irradiation | I need a draft of a letter written to an insurance company to appeal its decision to deny proton radiation therapy to Jane Doe, who is a 70 year-old woman with recurrent left breast invasive ductal carcinoma. Jane was first diagnosed on January 1st, 2023 with an UOQ left breast cT1cN0M0 IDC, ER+/PR+/Her2-. At that time she underwent a partial mastectomy that resected pathologic T1cN0M0 hormone receptor-positive, HER2-negative invasive ductal carcinoma. Thereafter she completed adjuvant whole breast radiation therapy to 40.05 Gy in 15 fractions. This was followed by adjuvant tamoxifen. However, a surveillance mammogram on January 1, 2024 identified a left breast upper outer quadrant 1.5 cm spiculated mass. On January 2, 2024 she underwent an ultrasound-guided core biopsy of this mass, and it identified recurrent grade 3 estrogen receptor-positive progesterone receptor-positive HER2-negative invasive ductal carcinoma. On January 3, 2024, Jane underwent a repeat lumpectomy and sentinel lymph node biopsy. Her cancer was excised with negative surgical margins and 0 of 2 lymph nodes were involved by cancer, pT1cN0M0. She now requires adjuvant radiation to decrease the risk of |

| | | | |
|---|---|---|---|
| | | | local recurrence. I have prescribed 42.56 CGE in 16 once-daily fractions. Explain that adjuvant radiation therapy with protons is indicated to protect OARs in the context of re-irradiation. Cite primary literature that proves that adjuvant proton therapy decreases the risk of damage to organs-at-risk compared to photon-based treatments. |
| Complex | Proton | Esophagus on trial (NRG GI-006) | Please provide me with a letter written to an insurance company that asks the insurer to approve reimbursement of a neoadjuvant proton radiation therapy course for John Doe, a 70 year-old man with esophageal squamous cell carcinoma who has enrolled on NRG GI-006, a national clinical trial: "Phase III Randomized Trial of Proton Beam Therapy (PBT) Versus Intensity Modulated Photon Radiotherapy (IMRT) for the Treatment of Esophageal Cancer." John is a 50 pack-year smoker and has averaged five alcoholic drinks per day for 20 years. He developed dysphagia to solid foods and lost 20 lb of weight in the course of two months. On January 1, 2024 he underwent an esophagogastroduodenoscopy, which visualized a mid-esophageal 3.5 cm ulcerating mass. Biopsy of this mass resulted as adenocarcinoma. FDG-PET/CT imaging on January 2, 2024 identified a single 1.5 cm lymph node at mediastinal station 8 with an SUV maximum of 10, and a mid-esophageal 3.5 cm FDG avid mass with an SUV maximum of 10. There was no evidence of distant metastatic disease. I have prescribed the patient 50.4 Gy in 28 fractions, and he will undergo treatment with concurrent carboplatin and paclitaxel under the direction of his medical oncologist. Explain in your letter that patients on this NRG-sponsored clinical trial requires upfront insurance approval of PBT at enrollment, irrespective of whether the patient is ultimately randomized to proton radiation. |

| | | | |
|---|---|---|---|
| Complex | Proton | Angiosarcoma | Write me a draft of a letter written to an medical insurance provider demanding it rescind denial of coverage for proton radiation therapy for Jane Doe, a 70 year-old female patient of mine who has a left breast radiation-induced angiosarcoma. Jane was first diagnosed with a left breast UOQ cT1cN0M0, ER+/PR+/Her2- IDC on January 1, 2024. At that time she underwent a partial mastectomy that resected pathologic T1c N0 M0 hormone receptor-positive HER2-negative invasive ductal carcinoma. Thereafter she completed adjuvant whole breast radiation therapy to 40.05 Gy in 15 fractions. This was followed by adjuvant tamoxifen. Unfortunately, on January 1, 2024 Jane presented to her oncologist for routine follow-up and was found to have a violaceous, papular, exophytic left breast mass. On January 2, 2024 this mass was biopsied and determined to be an angiosarcoma. The patient has already established care with a medical oncologist, Dr. Oncologist, who has informed the patient that she will likely need trimodality treatment with chemotherapy, surgery, and radiation therapy. Her prior radiation therapy delivered considerable dose to the brachial plexus, breast, lungs, and heart, among other organs. In order to mitigate dose deposition in these organs and consequent unacceptable toxicities, Jane Doe requires proton re-irradiation, not photon re-irradiation. Argue these details in your letter, and argue with medical literature why proton therapy is necessary for safe, efficacious treatment of her angiosarcoma. |
| Complex | Proton | Anus, young, Crohn's | I am a radiation oncologist, and I request that you draft an appeal letter written to a medical insurance company asking for coverage approval of IMPT for the definitive chemoradiation treatment of a cT2N1M0 locally advanced anal cancer in a 30-year-old man named John Doe who has a medical history of HIV (CD4: 200) and Crohn's disease (on vedolizumab). This treatment radiation treatment is delivered with curative intent but is delivered with concurrent 5-FU and mitomycin C chemotherapies and is associated with high rates of acute side effects including skin desquamation, cytopenias, and diarrhea. Late side effects of radiation therapy arise from scarring and healing and could include fistulas, ulceration, and bleeding. The risk of the side effects is greatly increased in this patient with Crohn's disease and marginally controlled HIV. The side effects of this treatment can be potentially life-threatening. The insurer refuses to cover treatment on the grounds that intensity modulated radiation therapy with photons is adequate treatment. The letter must convincingly counter that proton radiation is necessary to provide John Doe with the lowest probability of acute and late radiation side effects. It must argue that |

| | | | |
|---|---|---|---|
| | | | photon radiation, including IMRT, will not spare this young man's rectum and other organs as successfully as proton radiation. |
| Complex | Proton | Brain, young | Please write a letter to an insurance provider appealing its decision to deny 28 fractions of intensity modulated proton therapy (IMPT; total dose of 5040 CcGyE) as adjuvant treatment for a grade III 1p19q co-deleted right temporal lobe oligodendroglioma in a 30 year old man who is status-post gross total resection (1/1/24). Show why proton radiation therapy may decrease long-term, late effects of radiation in the healthy brain and surrounding tissues and potentially decrease neurocognitive decline compared to photon radiation. This is especially important in a young patient with a survival prognosis of many years. Support your reasoning with medical studies. |

| | | | |
|---|---|---|---|
| Complex | Proton | Generic | Please generate a generic letter template intended to appeal an insurance provider's rejection of medical coverage for proton radiotherapy. I need the letter to be easily adaptable to varying clinical scenarios in which proton radiation therapy may be indicated. I need a letter to generally explain the dosimetric advantages of proton radiation therapy over intensity modulated radiation therapy. I need this letter to cite primary literature, including clinical trials, that have shown instances in which proton radiation therapy engenders fewer or less severe toxicities to organs-at-risk than photon therapy. I need the letter to include guideline-directed indications for proton radiation therapy, such as in for radiation of children, reirradiation of previously irradiated sites, and radiation of patients with connective tissue diseases such as lupus or scleroderma. Ultimately, I need a well-reasoned, adaptable, data-supported letter template for the successful appeal of denial of insurance coverage for proton radiotherapy cases. |
| Complex | SBRT | Oligometastatic lung | I need a letter written to an insurance provider that requests reimbursement/coverage for stereotactic body radiation therapy (SBRT) to a single oligometastatic 3.5 cm right humeral metastasis in a patient with oligometastatic non-small-cell lung cancer. John Doe is a 70 year-old man with a history of coronary artery disease who was diagnosed with de novo metastatic lung adenocarcinoma after presenting to the emergency department on January 1, 2024 with a chief complaint of hemoptysis. A CT chest/abdomen/pelvis (January 2, 2024) identified a 3.5 cm right humeral lesion and a right peribronchial 3.5 cm spiculated right middle lobe lung mass with 4R and 4L mediastinal lymphadenopathy (largest node 3.5 cm). There was no radiographic evidence of disease elsewhere. On January 3, 2024, an endobronchial ultrasound and FNA of the 4R lymph node pathologically confirmed adenocarcinoma of the lung. He was staged as cT2N3M1. Next generation genomic sequencing did not identify any targetable mutation in the patient's cancer specimen. The patient was initiated on carboplatin, pemetrexed, and pembrolizumab on January 4, 2024. A surveillance CT CAP on January 5, 2024 visualized residual radiographic disease in the right humeral single metastatic site, which continued to grow, but no new metastatic lesions. Please explain to the insurance provider that this patient's overall survival is likely to be prolonged by aggressive consolidative SBRT to the right humeral lesion to 35 Gy in 5 once-daily fractions. This letter needs to refer to and cite supporting data from primary medical literature, including the SABR-COMET trial (Palma et al., J Clin Oncol, 2020). |

| | | | |
|---|---|---|---|
| Complex | SBRT | Oligoprogressive lung | I need a letter written to an insurance provider that requests reimbursement/coverage for stereotactic body radiation therapy (SBRT) to a single oligoprogressive 3.5 cm right humeral metastasis in a patient with initially widely metastatic non-small-cell lung cancer. John Doe is a 70 year-old man with a history of coronary artery disease who was diagnosed with de novo metastatic lung adenocarcinoma after presenting to the emergency department on January 1, 2024 with a chief complaint of hemoptysis. A CT chest/abdomen/pelvis (January 2, 2024) identified sclerotic and lytic lesions throughout the axial and appendicular skeleton, including a 3.5 cm right humeral lesion, and a right peribronchial 3.5 cm spiculated right middle lobe lung mass with 4R and 4L mediastinal lymphadenopathy (largest node 3.5 cm). On January 3, 2024, an endobronchial ultrasound and FNA of the 4R lymph node pathologically confirmed adenocarcinoma of the lung. He was staged as cT2N3M1. Next generation genomic sequencing did not identify any targetable mutation in the patient's cancer specimen. The patient was initiated on carboplatin, pemetrexed, and pembrolizumab on January 4, 2024. John's disease responded extremely well. A surveillance CT CAP on January 5, 2024 visualized interval resolution of almost all radiographic sites of disease, except the right humeral metastasis, which continued to grow. Please explain to the insurance provider that for this patient to remain on his current systemic therapy (which benefits John greatly by controlling all his disease with exception of the right humeral lesion) the right humeral lesion should be targeted aggressively with SBRT to 35 Gy in 5 once-daily fractions. This letter needs to refer to and cite supporting data from primary medical literature, including the CURB trial (Tsai et al., Lancet, 2023). |

| | | | |
|---|---|---|---|
| Complex | IGRT | 3D palliative re-irradiation | Explain to a medical insurer, in a formal appeal letter, why daily image-guided radiation therapy (IGRT) must be covered and reimbursed to safely re-treat a 3.5 cm right sacral prostate cancer metastasis with palliative 3D photon radiation therapy plan. Explain that this patient was diagnosed with de novo polymetastatic prostate cancer on January 1, 2024 after presenting to the emergency department with lower back pain and a PSA of 2000 ng/mL. After a sacral core biopsy pathologically confirmed prostate adenocarcinoma, the patient was initiated on androgen deprivation therapy and chemotherapy per the ARASCENS protocol (beginning January 2, 2024). On January 3, 2024, the patient completed palliative radiation therapy to the right sacrum to alleviate 7/10 constant right buttock and leg pain. The patient had complete pain relief from palliative radiation, but his pain has since recurred and CT scan (January 4, 2024) showed increase size of an osteoblastic right sacral lesion. I am prescribing him 30 Gy in 10 fractions palliative reirradiation to this site. Daily image guidance with each fraction is necessary to ensure proper alignment of our target and minimize overlap with previously irradiated organs, including the rectum and bladder. Explain all these details to the insurer in your letter, and reason why IGRT is medically necessary and therefore must be covered. |
